# WGS-Enabled Surveillance Improves Detection Of Transmission Events Within A Large Tertiary Care Hospital Trust In London

**DOI:** 10.64898/2026.03.24.26347804

**Authors:** Jonah Rodgus, Oskar Fraser-Krauss, Yalini Ravindra, Maria Getino, Ashleigh Myall, Chang Ho Yoon, Avishek Upadhya, Robert Peach, Sid Mookerjee, Alison Holmes, Elita Jauneikaite, Mauricio Barahona, Frances Davies

## Abstract

Infections caused by carbapenem-producing Enterobacterales (CPEs) are a persistent and growing threat in healthcare settings. Yet, current infection prevention and control (IPC) surveillance methods, which largely rely on the spatial and temporal proximity of patients, often misattribute or miss infection transmission events. Here, we develop and retrospectively evaluate an integrated methodology that combines analyses of ward-level patient movement data and whole-genome sequencing (WGS) data analyses, which provide measures of bacterial and plasmid similarity. Specifically, we evaluate this methodology across two datasets: a CPE outbreak of diverse carbapenem types (103 genomes, January 2021–March 2021) and an Imipenem-Hydrolysing β-lactamase-positive CPE outbreak (82 genomes, June 2016–October 2019), using standard clinical criteria and conservative genomic thresholds to quantify how often current IPC surveillance methods correctly identify genomically confirmed transmission events. Findings show that, across 3,423 patient contact–genome pairs, current IPC surveillance methods detected only 20.5% of genomically confirmed transmission events whilst maintaining 98.5% specificity, with missed events arising from temporal, spatial, and cross-species, mechanistic blindspots. In contrast, WGS-enabled IPC surveillance methods provided a 25–47-day earlier detection window and, in a linked economic evaluation, delivered annualised savings of up to £3.6 million, as well as a return on investment exceeding 2-fold in 7 of 8 cost scenarios. By operationalising high-throughput WGS data analysis with clinically relevant patient movement data, we evidence that it may be possible to disrupt and thereby mitigate the effects of AMR-driven CPE outbreaks, supporting investigations into the adoption of WGS-enabled IPC surveillance as a standard-of-care tool.

## Introduction

Current surveillance methods used by infection prevention and control (IPC) teams to detect transmission events underlying the spread of bacterial infections in hospitals largely rely on the spatial and temporal proximity of patients (Myall et al., 2022; Wan et al., 2024). Whilst useful as a baseline, these methods lack resolution, often leading to misattributed or missed infection transmission events (Sherry et al., 2022; Wan et al., 2024). When multiple transmission events are misattributed, missed, or detected too late, outbreaks may occur, resulting in significant downstream clinical and financial challenges (Otter et al., 2017). For example, healthcare-associated bacterial infections cost the NHS £2.7 billion annually and occupy 17% of hospital bed-days (Guest et al., 2019; Guest et al., 2020). As well as limited resolution in detecting single-species transmission events, current IPC surveillance methods are insensitive to cross-species transmission events mediated by mobile genetic elements, such as plasmids, which transfer important DNA material between different species of bacteria (Wan et al., 2024; Raabe et al., 2024; Sobkowiak et al., 2025). Compounding these challenges and thereby further complicating outbreak containment, the antimicrobial resistance (AMR) exhibited by many infection-causing bacterial species, such as carbapenem-producing Enterobacterales (CPEs), dramatically reduces the efficacy of key antimicrobial agents used to treat infected patients (Abbas et al., 2024; Nazir et al., 2025).

The integration of whole genome sequencing (WGS) data analyses into current IPC surveillance methods promises to address many of the challenges of outbreak containment, with outputs such as measures of genomic similarity between bacteria representing a more precise, higher resolution way to detect transmission events (Sundermann et al., 2022; Sundermann et al., 2024). Indeed, many view WGS-enabled IPC surveillance methods as the future of clinical surveillance (Jauneikaite et al, 2023), and evidence supporting such methods continues to grow. Recently, for example, Sundermann et al. (2025) reported a 3.2-fold return on investment (ROI) and approximately $700,000 in savings over two years by using WGS data analysis to guide early IPC interventions aimed at stopping transmission events. However, the routine integration of WGS data analysis into current IPC surveillance methods is constrained by imperfect technology infrastructure contributing to data silos, extended WGS turnaround times, and analysis complexity (Chindelevitch et al., 2023; Baker et al., 2023; Price et al., 2023).

In North-West London, the impact of WGS-enabled IPC surveillance methods has yet to be evaluated. Therefore, the availability of WGS data from CPEs of diverse carbapenemase types, including *Escherichia coli, Klebsiella pneumoniae, Citrobacter freundii*, and *Enterobacter hormaechei*, as well as Imipenem-Hydrolysing β-Lactamase (IMP)-positive CPEs from a large tertiary care hospital trust in London (Wan et al., 2024), presented a welcome opportunity to evaluate the value-add of WGS data analysis integration into current IPC surveillance methods. In this study, we evaluate WGS data analysis integration against three performance criteria: (i) timeliness, (ii) accuracy, and (iii) cost. Specifically, we hypothesised that (i) WGS data analysis integration enables the detection of transmission events up to 10 days earlier than current IPC surveillance methods; (ii) WGS data analysis integration improves transmission detection accuracy versus current IPC surveillance methods; and (iii), that WGS data analysis integration delivers a greater than 3-fold ROI, translating to over £1m in total annualised savings per hospital trust.

## Materials & Methods

### Whole-Genome Sequencing Data

#### Three-Month Sequential CPE Collection

The CPE Collection comprised genomes of 103 bacterial isolates from cross-infection screening swabs (n=98), urine samples (n=3), and soft tissue infections (n=2), positive for CPEs and non-invasive CPEs collected between 6th January 2021–27th March 2021 inclusive. These genomes carried diverse carbapenemase types, including OXA-48 (n=47), NDM (n=26), IMP (n=8), VIM (n=5), KPC (n=1), OXA-48 + NDM (n=2), and OXA-type (n=1). ***Three-Year Sequential IMP Collection***. The IMP Collection, described previously by Wan et al. (2024), comprised genomes of 82 IMP-positive CPEs from rectal swabs (n=77 isolates), urine samples (n=2), throat swabs (n=1), nasopharyngeal aspirate samples (n=1), and foot biopsy samples (n=1) collected between June 2016–October 2019.

### Genomic Data Handling

#### Quality Control

fastqc v0.12.1 (Andrews, 2010) and multiqc v0.4 (Ewels et al., 2016) were used to assess the quality of raw sequence reads. trimmomatic v0.39 (Bolger et al., 2014) was used to trim raw sequence reads. Reads were trimmed per specific instructions: to remove leading low-quality or N bases (below quality three); to remove trailing low-quality or N bases (below quality three); to scan the read with a 10-base wide sliding window, cutting when the average quality per base drops below 30, and to drop reads below 50 bases long. fastqc v0.12.1 and multiqc v0.4 were also used to assess read quality post-trimming. spades v3.15.5 (Bankevich et al., 2012) was used to assemble trimmed reads. quast v4.1 (Gurevich et al., 2013) was used to evaluate assembly quality before filtering. Assemblies were flagged as pass or fail depending on whether the total length, GC content, and N50 were within an appropriate range for each species (*K. pneumoniae*: Institut Pasteur, 2025; *E. coli*: Butters et al., 2025; *C. freundii*: Wan et al., 2020; *E. hormaechei*: Wee et al., 2021). bioawk v1.0 (Heng et al., 2017) was used to filter assemblies, keeping contigs ≥ 200 bp and with estimated coverage ≥ 2. quast v4.1 was also used to evaluate assembly quality post-filtering.

#### Species Identification

kraken2 v2.0.7-beta (Wood et at., 2019) and bracken v2.5.0 (Lu et al., 2017), along with a standard kraken2 bacteria database (last updated: 15th September 2023), were used to identify bacterial species from trimmed reads.

#### Whole Genome Similarity Comparisons

prokka v1.14.6 (Seemann et al., 2014) was used to annotate filtered assemblies. panaroo v1.5.2 (Tonkin-Hill et al., 2020) was used to construct a pangenome and identify core genes shared across genomes within each species. snp-dists v0.8.2 (Seemann, 2021) was used to calculate pairwise SNP distances from core genome alignments.

#### Plasmid Reconstruction & Characterisation

mobsuite v3.1.8 (Robertson & Nash, 2018) was used to reconstruct and characterise plasmid content from each filtered assembly.

#### Plasmid Similarity Comparisons

Given the fragmented nature of plasmid assemblies from the short-read WGS data, we retained for downstream plasmid similarity comparisons only the single largest contig from each plasmid assembly (typically representing the main backbone contig). pling v2.0.0 (Frolova et al., 2024) was used to calculate pairwise distances between plasmid backbones from both the same and different bacterial species. pling v2.0.0 quantifies plasmid similarity by comparing plasmid content across genomes using a network-based approach. sourmash v4.9.4 (Irber et al., 2024) was used for pre-filtering to reduce computational burden by excluding highly dissimilar plasmid pairs before pling analysis.

### Patient Data

#### Patient Screening

Detailed patient screening methods have been described previously by Wan et al. (2024).

#### Patient Movement Data

Pseudanonymised patient movement data were provided by the IPC team from the tertiary care hospital trust in accordance with our ethics. ***CPE Collection***. Patient-matched pseudanonymised movement data for the CPE Collection comprised 12,065 ward-day records from 89 unique patients, spanning a much larger time period, 21st August 2018–3rd February 2022, across four hospitals within a large tertiary care hospital trust in London. Movements were mapped to 40 distinct building codes and 87 distinct ward codes, providing a granular representation of inpatient locations over time. ***IMP Collection***. Patient-matched movement data, for the IMP Collection, comprised 7,207 ward-day records from 123 unique patients, captured between 8th June 2016–23rd October 2019. Movements were mapped to 16 distinct building codes and 79 distinct ward codes.

#### Ward Codes

A ward codes dictionary was developed to standardise ward identifiers across both pseudanonymised patient movement datasets to enable aggregation by clinical specialty. This dictionary comprises 166 unique ward codes (CPE Collection: 87; IMP Collection: 79) mapped to 26 standardised specialty categories. Ward codes were matched using four approaches: (1) direct matching against existing ward metadata (n=70); (2) department field extraction from IMP movement data (n=32); (3) fuzzy matching against ward code reference files (n=31); and (4) manual curation by clinical domain experts (n=33). The specialty categories with the most mapped ward codes were general medicine (n=28), general surgery (n=21), critical care (n=16), renal (n=13), and acute admissions (n=10).

### Temporal Filtering & Hierarchical Deduplication Of Contact–Genome Pairs

Genomes from both the CPE and IMP Collection were mapped to established patient contact events (**Figure 1**). To ensure epidemiological relevance, contact–genome pairs were filtered to include only those pairs where both genome collection dates occurred ≤90 days of the patient contact event. Contact–genome pairs were subsequently classified as either direct (same place, same time) or indirect (same place, different time). Further, to avoid double-counting, where genomes may have been simultaneously mapped to multiple patient contact events at different spatial levels (ward, building, and hospital), we used hierarchical deduplication such that each contact–genome pair appeared only once in the analysis dataset and that only the finest spatial resolution was kept per pair. Ward-level contacts were prioritised over building-level, which in turn were prioritised over hospital-level contacts.

**Figure 1.**
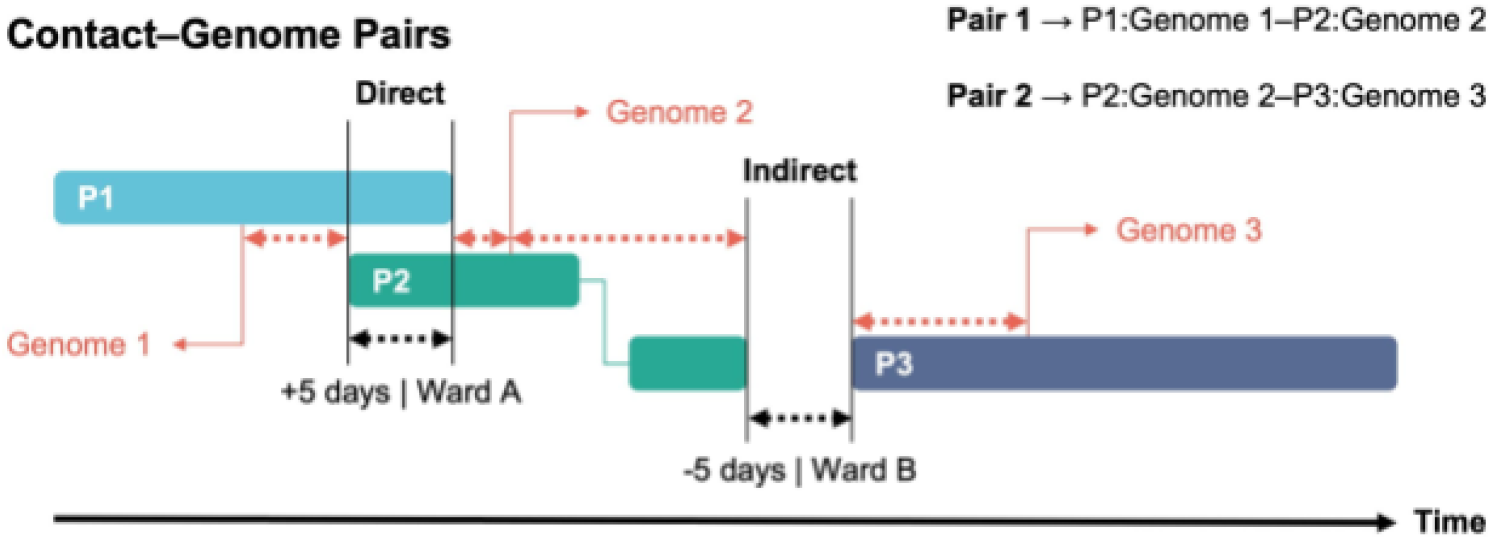
Genomes Mapped To Patient Contact Events. Schematic showing the mapping of genomes to direct and indirect patient contact events. P1 indicates patient P1; P2, patient 2; and P3, patient 3.

### Defining Clinical Criteria & Genomic Thresholds

We defined clinical criteria and genomic thresholds against which to test our hypotheses on (i) timeliness and (ii) accuracy, and ultimately evaluate the value-add of WGS data analysis integration. Clinical criteria constituted current IPC surveillance methods for identifying potential transmission events: that two patients present with an infection caused by the same species, in the same ward, ≤ 7 days apart, with at least one infection acquired post-admission. Specifically, the post-admission requirement distinguished potential ward-acquired infections from community importations. Genomic thresholds constituted single nucleotide polymorphism (SNP) distance ≤ 10 for within-species whole-genome comparisons and PLING distance = 0 for both within- and cross-species plasmid comparisons. **Supplementary Table 1** summarises patient contact events and corresponding genome-pair similarities. Both genomic distance thresholds were consistent with, but more conservative than, published thresholds for outbreak-related Enterobacterales (David et al., 2019; Ludden et al., 2020; Frolova et al., 2024; Raabe et al., 2024). Critically, such a low PLING distance captured identical plasmid backbone sharing indicative of recent horizontal gene transfer events, including cross-species plasmid transmission events that would have been neglected by whole-genome SNP-based approaches alone (Frolova et al., 2024).

### Evaluation Logic

To evaluate the clinical value-add of WGS data analysis integration against clinical criteria constituting current IPC surveillance methods, contact–genome pairs were classified into four categories. True positives (TPs): where a clinical action may have been triggered based on the acquisition of a new infection thought to have come from a patient contact event, and representative genomes were below the relatedness thresholds (reflecting confirmed transmission events; **Figure 2**). False positives (FPs): where a clinical action may have been triggered based on the acquisition of a new infection thought to have come from a patient contact event, but representative genomes were above the relatedness thresholds (reflecting misattributed transmission events). True negatives (TNs): where a clinical action may not have been triggered despite the acquisition of a new infection, and representative genomes were above the relatedness thresholds (reflecting correctly ignored, non-transmission patient contact events). False negatives (FNs): where clinical action may not have been triggered despite the acquisition of a new infection, but representative genomes were below the relatedness thresholds (reflecting missed transmission events).

**Figure 2.**
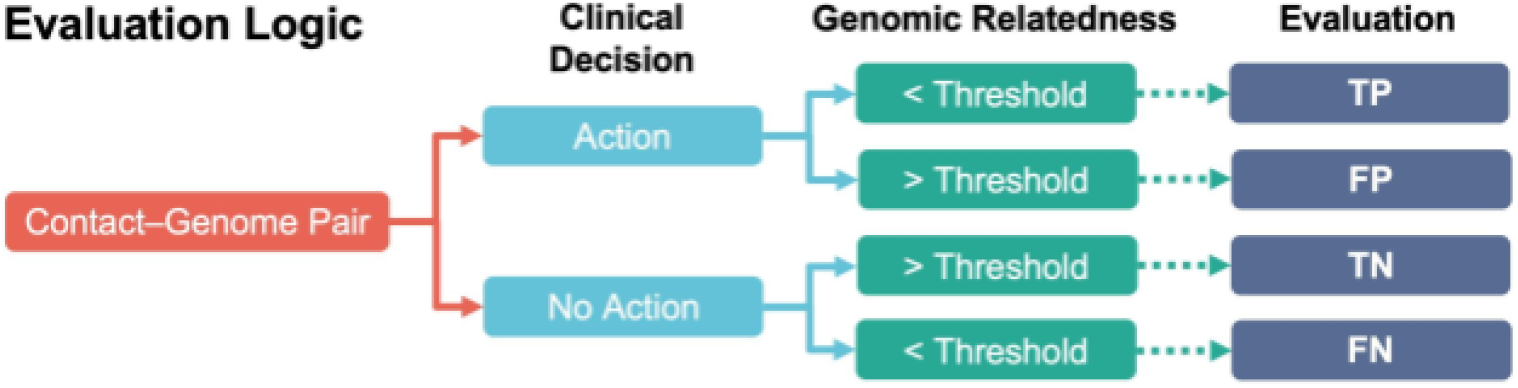
Logic For Retrospective Genomic Evaluation Of Clinical Decision-Making.

### Retrospective Evaluation Of Economic Impacts

#### Infection Cost Estimates

Cost estimates for new healthcare-associated infections were derived from published outbreak investigations, ranging from a conservative estimate of £3,288 per new infection based on averaged patient scenarios in England (Guest et al., 2020), to CPE-specific estimates of £23,375–£69,063 per new infection from outbreak scenarios in West London (Otter et al., 2017), Limerick (Dunne et al., 2019), and Paris (Atchade et al., 2022; **Supplementary Table 2**). ***WGS Cost Estimate***. A top-end WGS cost estimate of £100/genome was used. No bulk sequencing was assumed to be readily available. Therefore, the use of this estimate to calculate an ROI reflected a highly conservative approach.

#### Rule-Out Savings

For every unique misattributed (FP) transmission event flagged by existing clinical criteria but ruled out by WGS data analysis, we postulated that IPC teams could save money associated with escalating clinical action beyond just the treatment of a new infection. Such costs have been described exhaustively in a 2017 study on a 40-patient outbreak of CPEs that occurred across West London between July 2014–October 2015, and which led to costs of €1,100,000, comprising €312,000 actual material expenditure and €822,000 in opportunity costs (Otter et al., 2017; **Supplementary Table 2**). However, to be highly conservative in our evaluation, we factored in as a saving only the proportion of total costs reflecting actual material expenditure, not opportunity costs. ***Rule-In Savings***. For every unique (FN) transmission event missed by the clinical criteria but ruled in by WGS data analysis, we postulated that IPC teams could take clinical action to prevent new secondary infections reported to occur downstream of indexed CPE cases at a rate of 0.03 (Jamal et al., 2020; Park et al., 2020; **Supplementary Table 2**).

### Ethical Considerations

This study was carried out in accordance with ethics reference 21/LO/0170 (279677; protocol 21HH6538: “Investigation of Epidemiological and Pathogenic Factors Associated With Infectious Diseases”).

## Results

### Data Characterisation

The CPE Collection was collected from 89 distinct patients. Of these, 77 patients had single infection episodes, and 12 had multiple infection episodes (nine same species; three different species). Overall, there was an average of nine infection episodes per week across the 80-day study period between 6th January 2021–27th March 2021, with one genome collected per infection episode. The genomes represented 11 distinct species (*K. pneumoniae*, 40.8% [n=42 genomes]; *E. coli*, 23.3% [n=24]; *Enterobacter hormaechei*, 15.5% [n=16]; *C. freundii*, 8.74% [n=9]; *K. aerogenes*, 3.88% [n=4]; *K. variicola*, 1.94% [n=2]; *K. oxytoca*, 1.94% [n=2]; *K. michiganensis*, 0.97% [n=1]; *C. amalonaticus*, 0.97% [n=1]; *E. cloacae*, 0.97% [n=1], and *E. roggenkampii*, 0.97% [n=1]; **Figure 3**).

**Figure 3.**
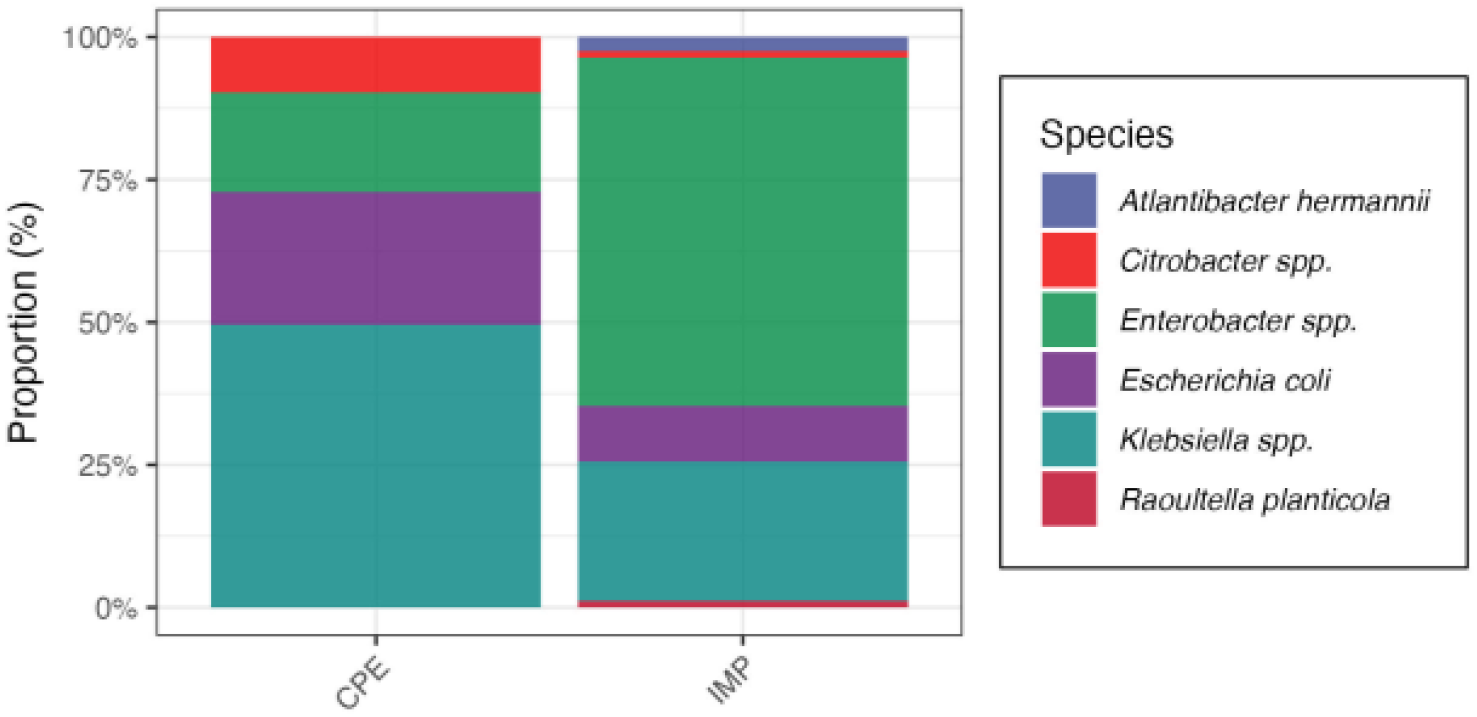
Species Breakdown For Each Genome Collection.

By comparison, the IMP Collection was collected over 1,000 days between June 2016–October 2019 from 78 distinct patients. Of these, 74 patients had single infection episodes, and four had multiple infection episodes (all different species). Overall, there was an average of 0.6 infection episodes per week across the study period. The genomes represented 15 distinct species (*E. hormaechei*, 42.7% [n=35]; *K. pneumoniae*, 18.3% [n=15]; *E. coli*, 9.76% [n=8]; *E. asburiae*, 7.32% [n=6]; *K. aerogenes*, 3.66% [n=3]; *E. cloacae*, 3.66% [n=3]; *E. kobei*, 2.44% [n=2]; *Atlantibacter hermannii*, 2.44% [n=2]; *E. ludwigii*, 2.44% [n=2]; *Roultella planticola*, 1.22% [n=1]; *K. quasipneumoniae*, 1.22% [n=1]; *E. bugandensis*, 1.22% [n=1]; *K. grimontii*, 1.22% [n=1]; *C. freundii*, 1.22% [n=1], and *E. roggenkampii*, 1.22% [n=1]; **Figure 1**).

After mapping, filtering, and hierarchical deduplication, the CPE Collection comprised 2,901 contact–genome pairs spanning four hospitals, 27 buildings, and 52 wards. By comparison, the IMP Collection comprised 522 contact–genome pairs spanning four hospitals, 12 buildings, and 25 wards.

### Retrospective Genomic Evaluation Of Clinical Decision-Making

For the CPE Collection, 62 contact–genome pairs involving 56 unique patients across 52 unique patient pairs met the clinical criteria for potential transmission events (same species infection, same ward, ≤ 7 days apart, with at least one infection acquired post-admission; **Supplementary Figure 1)**. Evaluating these clinical criteria across 2,901 contact–genome pairs (2,212 unique patient pairs), we observed a sensitivity of 29.7% (19/64) and specificity of 98.5% (2,794/2,837), with a positive predictive value (PPV) of 30.6% (19/62) and negative predictive value (NPV) of 98.4% (2,794/2,839; **Table 1**). At the patient pair level, these clinical criteria would have triggered investigation of 52 unique patient pairs, of which only 29% (15/52) had at least one genomically confirmed transmission event that justified a clinical action. The remaining 71% (37/52) of patient pairs showed no genomic relatedness across any of their genome comparisons, indicating that the triggering of a clinical action would have been an unnecessary waste of both time and resources.

**Table 1.** Confusion Matrix For CPE Collection.

|  | Contact–Genome Pairs |  | Total |
| --- | --- | --- | --- |
|  | < Genomic Threshold | > Genomic Threshold |  |
| IPC Action | 19 | 43 | 62 |
| No IPC Action | 45 | 2,794 | 2,839 |
| Total | 64 | 2,837 | 2,901 |

In total, WGS data analysis integration identified 45 FN contact–genome pairs (40 unique patient pairs) representing transmission events missed by the clinical criteria. These fell into three categories, deemed “blindspots”. First, temporal blindspots accounted for 24 same-species, same-ward contact–genome pairs where genomes were collected from infections > 7 days apart, with a mean interval of 25 days (median=20 days; maximum=76 days) between collections. Second, spatial blindspots accounted for 18 same-species contact–genome pairs from different wards, including four pairs from different buildings within the same hospital, and 14 pairs from different wards within the same building. Third, mechanistic blindspots accounted for three cross-species plasmid sharing events at the building (n=2) and hospital (n=1) level (7% [3/45] of FNs) that were invisible to existing methods, involving *E. coli*–*K. pneumoniae* (n=2) and *E. hormaechei*–*C. freundii* (n=1) pairs.

For the IMP Collection, 10 contact–genome pairs involving 16 unique patients across 10 unique patient pairs met the clinical criteria for potential transmission events (same species infection, same ward, ≤ 7 days apart, with at least one infection acquired post-admission). Evaluating these clinical criteria across 522 contact–genome pairs (463 unique patient pairs) revealed substantially lower sensitivity at 9.43% (5/53), though specificity remained high at 98.9% (464/469). The PPV was 50.0% (5/10), and the NPV was 90.6% (464/512; **Table 2**). Five FP clinical actions may have been triggered by the clinical criteria alone, where genomic data confirmed the strains were unrelated. Conversely, 48 FN contact–genome pairs were identified, representing missed transmission events.

**Table 2.** Confusion Matrix For IMP Collection.

|  | Contact–Genome Pairs |  | Total |
| --- | --- | --- | --- |
|  | < Genomic Threshold | > Genomic Threshold |  |
| IPC Action | 5 | 5 | 10 |
| No IPC Action | 48 | 464 | 512 |
| Total | 53 | 469 | 522 |

The IMP Collection exhibited a distinct distribution of surveillance blindspots compared to the CPE Collection. Temporal blindspots accounted for nine same-species, same-ward contact–genome pairs where genomes were collected from infections > 7 days apart, with a longer mean interval of 47 days (median=31; maximum=90). Spatial blindspots were more prominent, accounting for 15 same-species contact–genome pairs from different wards (48% [23/48] of all FNs), including 10 pairs from different buildings, and 5 pairs from different wards within the same building. Notably, mechanistic blindspots were more prevalent in the IMP Collection, with 23 cross-species plasmid sharing events detected. The most common species pairs involved *E. hormaechei*–*K. pneumoniae* (n=8), *K. aerogenes*–*E. hormaechei* (n=3), and *E. hormaechei*–*E. bugandensis* (n=2). Cross-species plasmid sharing events occurred at the hospital (n=13), building (n=6), and ward (n=4) levels (47.9% [23/48] of FNs).

Combining data for both collections, analysis across 3,423 contact–genome pairs yielded an overall sensitivity of 20.5% (24/117), specificity of 98.5% (3,258/3,306), PPV of 33.3% (24/72), and NPV of 97.2% (3,258/3,351; **Table 3**).

**Table 3.** Confusion Matrix Across Both Collections.

|  | Contact–Genome Pairs |  | Total |
| --- | --- | --- | --- |
|  | < Genomic Threshold | > Genomic Threshold |  |
| IPC Action | 24 | 48 | 72 |
| No IPC Action | 93 | 3,258 | 3,351 |
| Total | 117 | 3,306 | 3,423 |

### Retrospective Genomic Evaluation Of Economic Impacts

To evaluate the potential economic impact of integrating WGS data analysis into current IPC methods, we conducted a retrospective cost analysis comparing the expense of WGS integration against the costs associated with misattributed and missed transmission events. In brief, we distinguished between “rule out” savings, where WGS prevents unnecessary escalation of clinical action for genomically unrelated cases, and “rule in” savings, where WGS identifies otherwise-missed transmission events and enables prevention of secondary infections. A similar rules-based approach has been described previously by Parcell et al. (2021).

For the CPE Collection, we identified 37 FPs representing unique patient pairs for which a transmission event would have been misattributed but ruled out by WGS data analysis. Avoiding unnecessary clinical action in these cases yielded potential savings of £34,064–£715,493 depending on the cost model applied (**Table 4; Supplementary Table 3**). Additionally, WGS data analysis identified 40 FNs representing patient pairs for which a missed transmission event would have been ruled in by WGS data analysis. Appropriate clinical action here could prevent secondary infections costing between £3,946–£82,876. Combined, the total savings over the 80-day study period for the CPE Collection were £27,709–£788,068. When annualised, these savings translated to £126,424–£3,595,562 per year, representing an estimated ROI exceeding 2.69 (range: 2.69–76.51) relative to the £46,994 annualised WGS costs.

**Table 4.** Summary Of Retrospective Evaluation Of Economic Impacts.

| Collection | Cost Estimate | Reference | Rule Out Savings (£) | Rule In Savings (£) | Total Savings (£) | Annualised Savings (£) | Sequencing Costs (£) | Annualised Sequencing Costs (£) | ROI |
| --- | --- | --- | --- | --- | --- | --- | --- | --- | --- |
| **CPE** | Conservative (~£3,288) | Guest et al., 2020 | 34,064 | 3,946 | 27,709 | **126,424** | 10,300 | 46,994 | **2.69** |
| **CPE** | CPE-Specific Low (~£23,375) | Otter et al., 2017 | 242,165 | 28,050 | 259,915 | **1,185,862** | 10,300 | 46,994 | **25.23** |
| **CPE** | CPE-Specific Mid (~£44,074) | Dunne et al., 2019 | 456,607 | 52,889 | 499,195 | **2,277,579** | 10,300 | 46,994 | **48.47** |
| **CPE** | CPE-Specific High (~£69,063) | Atchade et al., 2022 | 715,493 | 82,876 | 788,068 | **3,595,562** | 10,300 | 46,994 | **76.51** |
| **IMP** | Conservative (~£3,288) | Guest et al., 2020 | 4,603 | 4,735 | 1,038 | **366** | 8,300 | 2,927 | **0.13** |
| **IMP** | CPE-Specific Low (~£23,375) | Otter et al., 2017 | 32,725 | 33,660 | 58,085 | **20,484** | 8,300 | 2,927 | **6.99** |
| **IMP** | CPE-Specific Mid (~£44,074) | Dunne et al., 2019 | 61,704 | 63,467 | 116,870 | **41,215** | 8,300 | 2,927 | **14.08** |
| **IMP** | CPE-Specific High (~£69,063) | Atchade et al., 2022 | 96,688 | 99,451 | 187,839 | **66,243** | 8,300 | 2,927 | **22.63** |

The IMP Collection analysis followed an analogous approach over the longer, 1,035-day study period. WGS data analysis ruled out 5 FP patient pairs for which a transmission event would have been misattributed but ruled out by WGS data analysis, generating savings of £4,603–£96,688 depending on the cost model applied (**Table 4; Supplementary Table 3**). A further 48 FN patient pairs for which a missed transmission event would have been ruled in by WGS data analysis were identified, with potential savings from prevented secondary infections estimated at £4,735–£99,451. Total savings for the IMP Collection were £1,038–£187,839. Given the extended study duration, annualisation yielded more modest annual savings of £366–£66,243, representing an estimated ROI of between 0.13–22.63 against the £2,927 annualised WGS costs.

## Discussion

### Clinical Decision-Making

Here, we show that current IPC surveillance methods for identifying infection transmission events, which require two patients present with an infection caused by the same species, in the same ward, ≤ 7 days apart, with at least one infection acquired post-admission, detect between 9–30% of genomically confirmed transmission events. Moreover, between 50–71% of clinical actions that may be triggered for patient pairs detected by current IPC surveillance methods appear unnecessary when evaluated against measures of genomic relatedness. Together, such findings provide substantial scope to enact, through WGS data integration, more precise interventions and reduce avoidable disruptions.

To this end, we show that for same-ward transmission events falling outside a 7-day time window, WGS-enabled IPC surveillance offers a mean early signal detection advantage of 25–47 days, addressing temporal blind spots in current IPC surveillance methods to create a sizeable opportunity for intervention before clusters evolve into large outbreaks. We also show that spatial blindspots are common, with genomically-linked cases frequently occurring in different wards, and sometimes different buildings, within the same hospital. Integrating WGS data analysis into IPC decision-making, therefore, supports more targeted, patient-pathways-focussed interventions at the building or hospital-level. Finally, we show that cross-species plasmid transmission account for between 7–48% of missed transmission events, representing a major mechanistic blindspot of current IPC surveillance methods. Detecting plasmid-mediated transmission events is therefore essential if hospital trusts are to properly contain outbreaks, rather than simply track where individual species occur.

### Economic Impacts

Using conservative assumptions, we show that WGS data analysis integration may deliver substantial net savings for the hospital trust studied, with positive annualised ROI estimates (well beyond 2-fold) generated for 7/8 cost models. At the lower bound of these positive ROI estimates, and excluding unquantified opportunity costs such as preserved bed capacity and averted elective surgery delays, the economic case for WGS-enabled IPC surveillance is strong. Indeed, total annualised savings ranged from £20,484–£3,595,561 for the cost models generating positive annualised ROI estimates. These findings align with those from a recent study on the real-world implementation of WGS-enabled IPC surveillance for a similarly sized outbreak in the US, that showed a 3.2-fold ROI and $700,000 in savings (Sundermann et al., 2025).

### Implementation

Beyond these primary clinical and economic impacts, this study also speaks to implementation. Routine use of WGS data analysis by IPC teams has so far been constrained by data siloes leading to limited linkage between operational IPC information and genomic outputs. By developing and applying a pipeline that integrates analysis of high-resolution patient movement data with WGS data analysis from clinically-derived genomes, we show that it is technically feasible to generate genomically informed, ward-, building-, and hospital-level views of transmission events, providing a practical foundation for WGS-enabled IPC surveillance aimed at containing outbreaks more effectively.

### Limitations

Chiefly, this study does not evaluate the operational feasibility of deploying WGS-enabled IPC surveillance in a true real-time clinical environment. Our analyses were conducted retrospectively, meaning system latency that results from sequencing turnaround constraints, and integration with IPC team workflows were not formally assessed. Consequently, whilst epidemiological performance can be inferred, the practical challenges associated with live deployment remain unquantified. Secondly, our findings derive from data within a single NHS Trust, limiting their generalisability. Hospital infrastructure, ward connectivity, patient case-mix, admission pathways, screening policies, and local epidemiology of CPEs and specific CPE resistance mechanisms, vary substantially across geographies and institutions. Thirdly, the patient movement data to which we had access were incomplete at the ward level for a non-negligible subset of genomes, meaning that not all patient-location transitions could be reconstructed at the highest resolution. This may have led to underestimation of contact events detected per current IPC surveillance methods, particularly during short-duration ward overlaps or inter-ward transfers occurring outside available timestamps.

Additionally, our pipeline for defining genomic relatedness relied on fixed thresholds that represent pragmatic but perhaps oversimplified approximations of biological reality. The SNP distance and PLING distance thresholds applied in this study were intentionally more conservative than, but consistent with, prior work (David et al., 2019; Ludden et al., 2020; Frolova et al., 2024), but we did not perform sensitivity analyses exploring a range of alternative thresholds. This limits inferences about the robustness of our findings to more or less stringent distance cut-offs. Fixed thresholds can also be problematic for inferring transmission in noisy genomic datasets, where relatedness may be better represented probabilistically (Stimson et al., 2019). Similarly, whilst we incorporated plasmid-similarity measures to capture potential cross-species transmission, our handling of short-read WGS data was inherently constrained. Here, we retained only the largest contig per plasmid assembly when calculating PLING distances, which may not have fully represented multipart plasmids or the broader plasmidome diversity present. Consequently, this approach may have underestimated plasmid diversity and overlooked more complex plasmid-mediated transmission events. Recent work has demonstrated that cross-species plasmid transmission events can be more readily identified by using long-read sequencing (Sobkowiak et al., 2025).

Finally, our cost evaluation methodology presents several limitations. Cost estimates were derived from published outbreak investigations rather than hospital-specific financial data, meaning the absolute savings and ROI values reported here may not directly reflect the trust’s true cost base. Additionally, some of the reference values we used to inform our analysis may be outdated relative to current NHS cost structures and operational expenditures. Our analysis also assumed the availability of pre-existing capacity for isolate preparation and downstream genomic analysis, such as existing wet-lab and bioinformatics resourcing, and therefore excluded associated staffing and infrastructure costs. Other studies have provided a detailed breakdown of per-isolate WGS costs when considered in addition to equipment, consumables, staffing, and batch size (Alleweldt et al., 2021). As a result, the evaluation of economic impacts presented likely represents an optimistic lower bound on implementation costs and an upper bound on potential ROI.

### Further Work

Future work should focus on addressing the technical and operational challenges associated with implementing near-real-time WGS-enabled IPC surveillance in a live clinical environment; specifically, to establish a system for rapid, preferably long-read WGS data ingestion and downstream analysis capable of producing actionable intelligence for IPC teams. Key questions include how such a system would perform relative to live IPC surveillance methods, both with and without existing proprietary analytics platforms, and whether the performance demonstrated retrospectively in this study can be maintained under live clinical constraints. If such performance is demonstrated, hospitals should then push to deploy WGS-enabled IPC surveillance alert systems. Moreover, national health systems should establish governance, infrastructure, and reporting standards to support the scalable adoption of such alert systems.

## Conclusion

Here, we show that integrating WGS data analysis into current IPC surveillance methods enables more timely, accurate, and cost-effective detection of transmission events that cause the spread of bacterial infections in hospitals. By operationalising high-throughput WGS data analysis with clinically relevant patient movement data analysis, it may be possible to disrupt and thereby mitigate the effects of AMR-driven CPE outbreaks, supporting urgent investigations into the adoption of WGS-enabled IPC surveillance in live clinical environments as a standard-of-care tool.

## Data Availability

All data produced in the present study are available upon reasonable request to the authors.

## Supplementary Figures

**Supplementary Figure 1.**
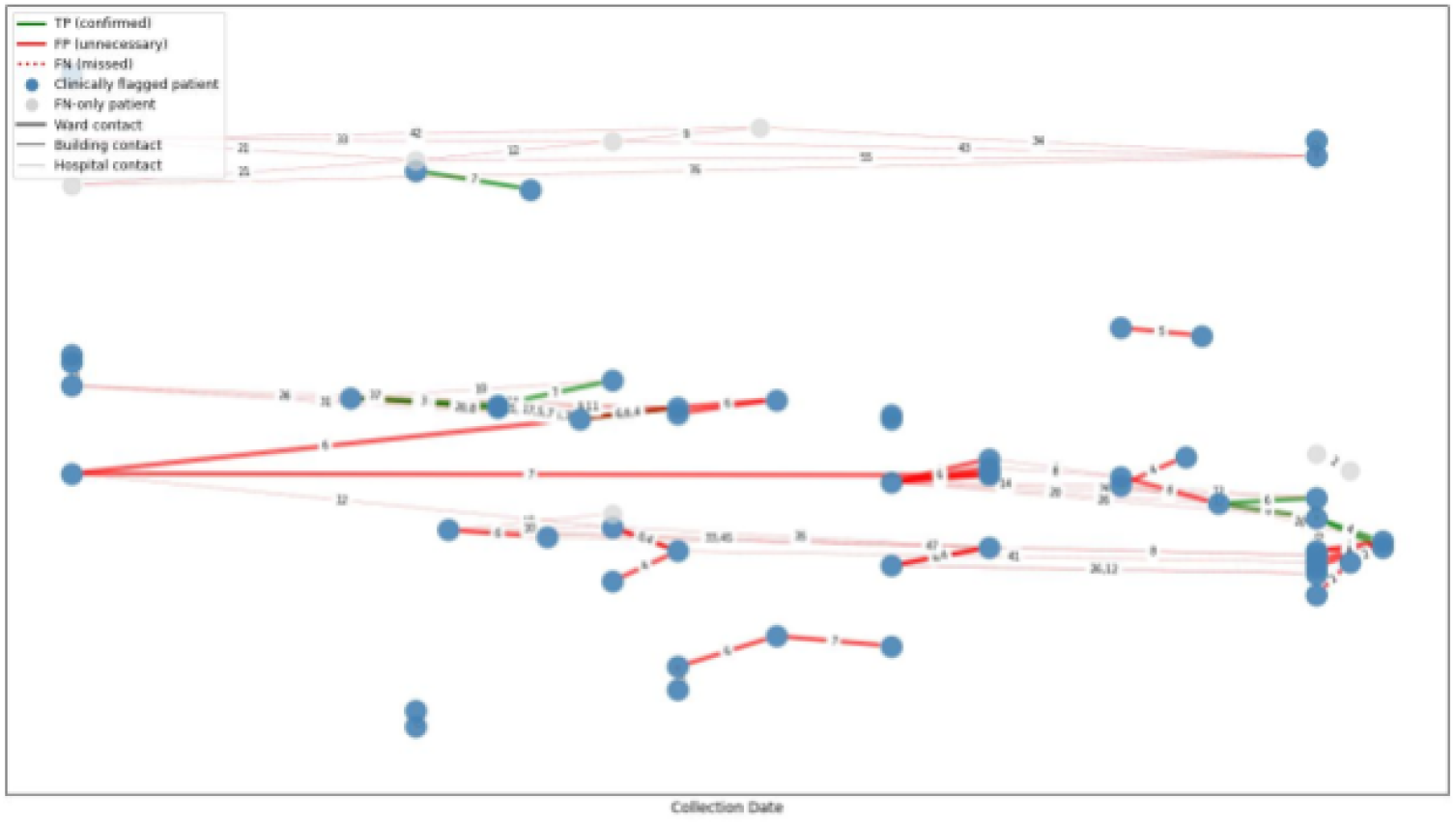
Patient transmission network for the ICHNT CPE Collection. Each node represents a patient and each edge a genome pair comparison. Blue nodes indicate patients involved in at least one clinically flagged pair. Grey nodes indicate patients involved only in genomically detected (false negative; FN) pairs. Green edges indicate true positives (TP; clinical action confirmed by genomic data); red solid edges indicate false positives (FP; clinical action not supported by genomic data). Red dotted edges indicate false negatives (FN; genomically related pairs not flagged by clinical criteria). Edge thickness indicates spatial proximity of contact (ward > building > hospital). Numbers on edges indicate days between sample collection dates. X-axis represents each earliest isolate collection date for each patient.

